# Test-retest reliability of electronic hand dynamometer in healthy adults- A cross sectional study

**DOI:** 10.1101/2025.08.14.25333572

**Authors:** Amal K Francis, H A Kalashree, Jakson K. Joseph

## Abstract

**Introduction:** Hand grip strength (HGS), is a crucial indicator of muscle function and general physical capacity of the elderly. Few tools are available to measure grip strength, including pneumatic instruments, spring devices, and hydraulic systems. However, among these, hydraulic dynamometers, such as the Jamar dynamometer, have been considered the gold standard due to their superior reliability and precision. Despite their advantages, these tools often come with limitations, such as high cost. Hence, the purpose of this study is to estimate the test-retest reliability of the electronic hand dynamometer for grip-strength measurement and also to compare the reliability of the electronic hand dynamometer in flexed and extended elbow.

**Methods:** The participants were recruited via convenience sampling and this ross sectional study followed the procedure as per the American Society of Hand Therapist (ASHT) recommended standardized position for grip strength.

**Results:** The reliability of the electronic hand dynamometer for the right side with the elbow flexed was found to be 0.94 while for left side, it was found to be 0.91. Meanwhile, the reliability for the right side with the elbow extended was found to be 0.98, and for the left side, it was 0.97.

**Conclusion:** The findings of this study demonstrate that the Camry electronic hand dynamometer has excellent reliability in testing grip strength. The grip strength was found to produce maximum strength when measured in an elbow extended position.

## Introduction

Hand grip strength (HGS) is a crucial indicator of muscle function and general physical capacity of the elderly. HGS has a significant relationship with age-related loss of muscle mass, strength, function, and sarcopenia. There have also been reports that it can reveal a person’s health ^[1].^

Grip strength is a vital indicator of overall physical health and an important parameter in clinical and rehabilitation settings. Physical therapists frequently assess grip strength to establish a baseline hand grip strength, evaluate treatment outcomes, and useful to monitor recovery from conditions affecting the hand function and also it is a prognostic factor for post operative prognostic factor ^[2-4]^

Reliable and valid measurements of grip strength are critical to ensure reproducibility and accuracy across repeated evaluations, which are typically enhanced through standardized assessment protocols recommended by organizations such as the American Society of Hand Therapists ^[5-6]^.

Several tools are available to measure grip strength, including pneumatic instruments, spring devices, and hydraulic systems. However, among these, hydraulic dynamometers, such as the Jamar dynamometer, have been considered the gold standard due to their superior reliability and precision. Despite their advantages, these tools often come with limitations, such as high cost and lack of advanced features for data management such as data storage and processing capabilities that allow for precise measurement and tracking of force exerted by users. Which is essential for assessing muscle strength and rehabilitation progress ^[6-8].^

Recently, researchers have come up with an instrument, Camry electronic hand dynamometer, to measure maximal isometric grip strength. Added advantage of this Camry electronic hand dynamometer (EH101) is, it can auto capture maximum achieved grip strength which can be calibrated to 200 lbs (90 kg) and display the value ^[9-11].^ It can assess results according to age and gender as well as compare current records to the last one. Since the values are auto-captured within the instrument, the examiner need not keep up his/her stationary position. The instrument has adjustable grip sizes to account for varying hand sizes. It has a digital LCD read-out, with on-screen rating of results according to age and gender. the instrument saves and stores results for fast retrieval with up to 19 users ^[11].^ Moreover, it is economical in comparison to study instrument like Jamar and Baseline. Grip dynamometer measures the maximum voluntary effort by the participant ^[12]^. Hence, even though the instrument is precise, internal threats to validity may persist. Hence it is necessary to assess test-retest stability of the measurement to control for its test trial variability. The manufacturer has recommended two positions of testing. The more reliable position must be determined.

Reliability studies of Camry electronic hand dynamometer (EH101) showing optimal positions of arm have not been conducted. Hence the purpose of this study is to estimate the test-retest reliability of Camry electronic hand dynamometer measurement and also to compare reliability of the reliability of electronic hand dynamometer in various positions of both sides, i.e., flexed and extended elbow.

## Methods

This cross-sectional study design aims to measure the grip strength using Camry electronic hand dynamometer (EH101) and to assess the reliability of the same and recruited the samples via convenient sampling method from JSS College of physiotherapy. The sample size was estimated using the following formula and 19 participants were recruited in each age group.

Formula:

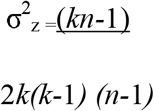

The duration of the study is 1 year and participants included were Normal adults of 17 years of age and above with both male and female via convenience sampling method. However, adults with any past history of hand injury, neurological deficits, visual or auditory dysfunctions, any joint disease, any previous history of fracture, upper limb pain syndrome and subjects with cervical pathology were excluded from the study.

Institutional Ethics Committee (IEC) was obtained (JSS/MC/IEC/02/664/2015-16), written consent was obtained from the participants before the commencement of the study, and materials used for the study include Camry Electronic Hand Dynamometer, a standard Chair (seat height: 45 cm, backrest height: 90 cm), Pen, and paper to note the data.

At the beginning of the study, demographic data (age, sex, weight, height, dominant hand) was obtained and recorded. The participants were explained about the procedure and Rater,(a licenced physiotherapist and post graduate student)demonstrated the procedure as per the American Society of Hand Therapists (ASHT) recommended standardized position for grip strength and collected the data^[9]^.

Sitting position in a straight-back chair with both feet flat on the floor was the starting position. The dominant hand was placed on the ipsilateral thigh, shoulder adducted to zero degree and neutral rotation. Hand grip strength was measured with elbow at a right angle to the shoulder and elbow in the extended position with arm hanging by the side ^[9]^. The handle of the dynamometer was adjusted with the base resting on the metacarpal (heel of palm). Before the participants asked to asked to perform maximum hand grip, the procedure of maximum handgrip strength was demonstrated by the rater. The participants were asked to squeeze the dynamometer with maximum isometric effort, which was maintained for about 5 seconds. No other body movement was allowed. Participants were asked to practice before the actual test commencement, followed by a 5-minute rest period ^[9]^.

To determine test-retest reliability, each participant underwent two testing sessions on consecutive days at the same time of day and strictly followed the ASHT guidelines including participants position, dynamometer settings and verbal instructions. Participants were asked to perform 3 attempts with a rest period of 30 seconds between each attempt. No feedback regarding performance was given however, the participants were encouraged consistently by instructing the participants to “squeeze harder … harder” ^[9]^. The reading of the three trials was auto recorded in the dynamometer in kgs, and mean was taken to minimize the fatigue bias. A retest was done the next day at the same time with the same procedure, and the mean of the retest readings was taken. Before the attempt, warm-up was before 15 minutes of testing ^[9].^

Insert image 1 and 2

## Results

Data were collected over six months. Dropouts were recorded, but no participants withdrew. Statistical analysis was performed using SPSS v23. A total of 114 participants were included in this study. The included samples were divided into six groups according to age, with 19 samples in each group that is Age groups:17-26 years,27-36 years,37-46 years,47-56 years,57-66 years,> 66 years data analysis was done using SPSS software 23.0 was used for statistical analysis. The mean and standard deviation were calculated. Pearson’s correlation was used to find the correlation between the best of two of the three trials, which were considered and calculated to evaluate the degree of agreement of participants’ scores on both occasions. SEM was used to evaluate the measurement precision and convey information about the magnitude of measurement. The mean and standard deviation results are presented in Table 1. Reliability is expressed by the correlation coefficient, which ranges from 0 to 1, with 0 showing no reliability and 1 showing perfect reliability ^[10]^. ICC was found to be 0.95. The reliability of the electronic hand dynamometer for the right side with the elbow flexed was found to be 0.94, while for the left side, it was found to be 0.91. Meanwhile, the reliability for the right side with elbow extended was found to be 0.98, and for the left side was 0.97. The high ICC values (ranging from 0.91 to 0.98) indicate excellent reliability, suggesting minimal bias between test and retest measurements. SEM values are listed in Table 2 below.

**Table 1:** Showing Mean and Standard deviation of grip strength measurement

|  | N | Minimum(kgs) | Maximum(kgs) | Mean(kgs) | Std.Deviation(kgs) |
| --- | --- | --- | --- | --- | --- |
| EFR1 | 230 | 9.10 | 54.50 | 27.90 | 10.28 |
| EFRR1 | 228 | 11.00 | 54.30 | 27.76 | 10.07 |
| EER1 | 228 | 10.70 | 54.20 | 27.47 | 10.10 |
| EERR1 | 228 | 10.20 | 53.20 | 26.80 | 9.96 |
| EFL1 | 228 | 3.20 | 50.10 | 24.66 | 9.22 |
| EFLR1 | 228 | 9.90 | 51.10 | 23.94 | 8.88 |
| EEL1 | 228 | 9.10 | 48.20 | 24.01 | 8.85 |
| EELR1 | 228 | 9.10 | 50.10 | 23.33 | 8.37 |

*Abbreviations:* N- Total number of participants, EFR1-Elbow flexed right test1, EFRR1-Elbow flexed right retest, EER1-Elbow extended right test1, EERR1-Elbow extended right retest,
EFL1-Elbow flexed left test1, EFLR1-Elbow flexed left retest, EEL1-Elbow extended left test1, EELR1-Elbow extended left retest
Note: Mean and standard deviation of the two trials considered

**Table 2:** Showing reliability results.

|  |  | Coefficient of variance | Standard error of measurement | Interclass correlation coefficient | Test –retest reliability |  |
| --- | --- | --- | --- | --- | --- | --- |
|  | EFR1 | 36.84 | 2.30 |  | REF | .94 |
| RIGHT | EFRR1 | 36.27 | 2.25 | .95 | REE | .98 |
|  | EER1 | 36.76 | 2.26 |  |  |  |
|  | EERR1 | 37.16 | 2.23 |  |  |  |
|  | EFL1 | 37.38 | 2.06 |  | LEF | .91 |
| LEFT | EFLR1 | 37.09 | 1.98 |  | LEE | .97 |
|  | EEL1 | 36.85 | 1.98 |  |  |  |
|  | EELR1 | 35.87 | 1.87 |  |  |  |

Abbreviations: EFR1-Elbow flexed right test1,EFRR1-Elbow flexed right retest,EER1-Elbow extended right test1,EERR1-Elbow extended right retest ,EFL1-Elbow flexed left test1,EFLR1-Elbow flexed left retest,EEL1-Elbow extended left test1,EELR1-Elbow extended left retest,REF – Right Elbow Flexed,REE – Right Elbow Extended,LEF – Left Elbow Flexed and LEE – Left Elbow Extended

## Discussion

The current study aims to estimate the test-retest reliability of electronic hand dynamometer for grip-strength measurement and also to compare reliability of the reliability of electronic hand dynamometer in various positions of both sides, i.e., flexed and extended elbow and the results of the study reveals excellent test-retest reliability of electronic hand dynamometer in healthy adults with ICC of 0.95 which indicates high degree of consistency measurement between two testing occasions with an interval of 1 day. The instrument used was Camry electronic hand dynamometer. This instrument provided stable and similar measurements at the two occasions on which the grip strength was tested. There are several studies have investigated the effect of various body positions on grip strength measurements. In this study the position used was sitting upright on a chair. This position is found to be the maximum force generating posture ^[12]^.

Three trials were performed on both the occasions of strength testing. The mean of the three trials were taken for the strength measurements to be accurate. Out of the three trials, first two were considered after correlation since there were chances errors with the correlation of the third trial due to muscular fatigue.^[8]^ The measurement procedures were performed with elbow in two positions: flexed and extended. With an interval of one day between the two testing occasions, the ICC was found to be 0.95 which is considered to be excellent. All participants included were right-hand dominant. The reliable measurement of the electronic hand dynamometer was indicated by the high value of the ICC score.

The highest mean grip-strength measurement in the present study was found when the elbow joint was placed in 0 degree of flexion i.e. fully extended position as compared to elbow flexed at 90 degrees^[11]^. The reliability of the instrument was found to be better in the elbow extended position than the elbow flexed position with a score of 0.98 for the extended position.

Participants, too felt the fully extended position was easier for them to perform the action. Kuzala et al did a study on grip strength measurement in elbow flexed and extended position and found better scores of grip strength in the elbow extended position than the elbow at 30, 45, 90 and 135 degrees of flexion.^[12].^

When the elbow joint is in a flexed position, the angular variations during the procedures may affect the test-retest reliability. Whereas in the elbow extended position, i.e. the close pack position of elbow, the joint gains maximum stability and thus the measurement errors can be minimized also gives good reliability. ^[13-14]^.

As compared to the standard jamar hand dynamometer, the electronic hand dynamometer is economically cheaper and easily available in market. The procedure to check grip strength using this equipment is also very easy.^[15]^. Thus, it complies with the patients’ need to assess the grip strength at home as well as for the clinicians. The internal memory storage of the dynamometer can store the past grip strength values in the machine. This can help patient know about the progress in grip strength after each exercise session. Thus, Camry electronic hand-held dynamometer can be used for assessing grip strength in healthy adults of both genders and all ages.^[15]^. The strength of the study is test-retest reliability of the electronic hand dynamometer showed excellent reliability and the scores were the best when the elbow was held in fully extended position during the testing procedure and fatigue was minimized which reduced the errors and Camry electronic hand dynamometer has excellent intra-rater reliability. ^[15-16]^. The clinical implication of the current study describes grip strength is one the important indicator of hand function and risk of falls hence measuring grip strength clinically is considered has an important evaluation and since the Camry electronic hand held dynamometer is cost effective, portable, requires less time, and needs minimal no experience to use this tool and reliable instrument for grip strength measurement among healthy adults of all age groups as hence it can be used in clinical practice. However, the limitation of the study is obtained results cannot be generalized to the general population and the sample of the current study was homogeneous in age distribution and activity level hence no subgroup analyses conducted; future research should stratify analyses by gender, include anthropometric measures and assess additional physical parameters to understand their independent effects on grip strength.

## Conclusion

The findings of this study demonstrate that the Camry electronic hand dynamometer (EH101) demonstrated excellent test–retest reliability, with high ICC values indicating consistent strength measurements across sessions. The grip strength was found to produce maximum strength when measured in an elbow extended position.

## Ethical approval

Ethical approval for this study was obtained from the Institutional Ethics Committee of JSS Medical College (Approval No: JSS/MC/IEC/02/664/2015-16) prior to commencement of the research and data collection was completed within the original ethics approval period.

## Data Availability

The datasets generated and/or analyzed during the current study are available from the corresponding author on request.

## Acknowledgement

Dr.Kavitha raja, Principal JSS College of Physiotherapy for being very supportive in all the aspects to complete the research

## Authors contribution

1. Concept and design of study or acquisition of data or analyses and interpretation of data-Amal K Francis1
2. Drafting the article or revising it critically for important intellectual content-Kalashree H A2
3. Final approval of the version to be published-Jakson K Joseph3

